# Characterisation of Microbial Community Dynamics and Antibiotic Resistance Gene Dissemination in Malaysian Wastewater during the COVID-19 Pandemic

**DOI:** 10.1101/2024.07.25.24311021

**Authors:** Umama Shahid, Hooi Suet Li, Lim Shu Yong, Alijah Mohd Aris, Khor Bee Chin, Qasim Ayub, Tan Hock Siew

## Abstract

Wastewater is a well-known hotspot for pathogens and spread of antibiotic resistance across species. Surveillance of wastewater microbial community can help draw clearer representation of actively culturing taxonomic groups and resistance-inducing mobile genetic elements before and after treatment. Studies have suggested that COVID-19 pandemic may also have caused increased dissemination of antibiotic-resistance genes (ARGs) and antibiotic-resistant bacteria in wastewater. Although immensely significant, no research has yet been performed on Malaysian wastewater microbial community and ARGs or their correlation with COVID-19 infections. This study utilised 16S metagenomics approach to characterise microbial community in Malaysian wastewater during high and low-case phases of pandemic. Among 20 most prevalent genera around Kuala Lumpur, Malaysia, those belonging to Bacteriodales, Bacillales, Actinomycetales and opportunistic pathogens-Arcobacters, Flavobacteria, and Campylobacterales, Neisseriales, were enriched during high-case periods of the COVID-19 pandemic. Copy number profiling of ARGs in water samples showed prevalence of elements conferring resistance to antibiotics like sulphonamides, cephalosporins, and colistin. High prevalence of *intI1* and other ion-based transporters in samples highlight an extensive risk of horizontal gene transfer to previously susceptible species. Our study emphasises the importance of wastewater surveillance in understanding microbial community dynamics and ARG dissemination, particularly during public health crises like the COVID-19 pandemic.

## Introduction

Wastewater is the water generated by households, industries or agricultural runoffs. It contains ample nutrients, heavy metals, and organic and inorganic compounds, which offer a favourable environment for microbial growth, including pathogens (Chahal et al. 2016). Wastewater treatment plants (WWTPs) are built to treat this water before discharging it into the environment. The major bacterial phyla in WWTP are Proteobacteria, Bacteroidetes, and Firmicutes (Numberger et al. 2019). Although these species are consistent, a variation in bacterial genera is often observed as the wastewater composition constantly shapes the microbial profile in WWTP. Several investigations have shown prevalence of *Clostridium* species (spp.), *E. coli, Enterococcus* spp., *Salmonella* spp., *Shigella* spp., *Vibrio cholerae, Yersinia* spp., *Pseudomonas* spp. and *Legionella pneumophil*. A study observed that even after treatment, some pathogens, such as *E. coli*, could persist in the effluent, thus posing possible risks to the surrounding community (Shannon et al. 2007). Several studies have suggested that the COVID-19 pandemic could have led to a spike in the dissemination rate of antibiotic resistance genes (ARGs) and antibiotic-resistant bacteria (ARB) in the environment, mainly due to the frequent usage of personal hygiene products, disinfectants and antibiotics (Rezasoltani et al. 2020; Wang et al. 2022; Hu et al. 2023). Hence, identifying pathogens is now even more crucial for effective wastewater treatment.

## Material and Methods

### Study area

We characterised the wastewater microbial community around Klang Valley in the Federal Territory of Kuala Lumpur amidst the COVID-19 outbreak from October 2020 to February 2021 using partial 16S rRNA (V3-V4) sequencing. Additionally, we evaluated the effect of high (∼1780) and low incidence (∼ 167 cases) of COVID-19 infections during this period on the microbial community and ARGs around the studied Klang Valley area.

### Sample collection

Samples were collected from inlet points of two WWTPs (A and B) that are 15km apart from each other in the Klang Valley that serve a combined population of about 0.27 million. All samples (untreated wastewater) were collected in 1L sterile Schott bottles at their respective locations and were stored at 4°C until further processing. To inactivate viral particles, the samples were exposed to ultraviolet (UV) light for 20 minutes in a biosafety cabinet (BSC) before incubating in a water bath at 60°C for 90 minutes. Subsequently, for each sample, 500mL of wastewater was filtered through 0.20 µm nylon membrane filters (Merck Millipore, Burlington, MA, USA) by vacuum filtration. Later, the membrane filters were cut into tiny pieces and transferred into a PowerBead tube of the DNeasy PowerSoil Kit (Qiagen, Hilden, Germany), followed by DNA extraction per the manufacturer’s protocol.

### 16S rRNA gene sequencing

The DNA samples were used to amplify the V3-V4 region of the 16S rRNA gene. The forward primer was designed with Illumina 5’ overhang adapter sequence followed by binding site sequence (5’-CCTACGGGNGGCWGCAG-3’), whereas the reverse primer was designed with another 5’overhang adapter followed by binding site sequence (5’-GACTACHVGGGTATCTAATCC-3’). For each sample, the amplification was performed twice to obtain technical replicates. The libraries thus constructed were then sequenced using the Illumina Miseq platform (Illumina, San Diego, CA, USA) with 2 × 250 bp generating 100,000 reads per sample.

### Data Processing and Analysis

All the analyses were performed on the Easy Microbiome Analysis Platform (EasyMAP), which incorporates modules from QIIME2, Linear Discriminant Analysis Effect Size (LefSe), and Phylogenetic Investigation of Communities by Reconstruction of Unobserved States (PICRUSt) (Hung et al., 2021). Briefly, QIIME2 plug-in q2-dada2 was used to perform the quality control of the raw sequences, which included pre-processing to remove primer tags and low-quality sequences and merging paired ends, followed by Operational Taxonomic Units (OTU) table construction. Alpha diversity was evaluated in terms of the quantitative richness and evenness of the community (Shannon diversity), the qualitative richness of the community (observed OTUs), phylogenetic relationships between the features (Faith Phylogenetic Diversity), and community evenness (Peilou’s Evenness). Beta diversity check included a qualitative measure of community dissimilarity (Jaccard distance), a quantitative measure of community dissimilarity (Bray-Curtis distance), qualitative phylogenetic dissimilarities between features (unweighted UniFrac distance), and quantitative phylogenetic dissimilarities between features (weighted UniFrac). The taxonomic assignment was performed using the Greengenes V34 classifier with default parameters. To visualise the distribution of abundant OTUs, a heatmap was generated to visualise the 32 most abundant taxa across all 20 samples. The differentially abundant taxa were identified using LEfSe (α=0.05, Wilcoxon test) with LDA score 2 and Kruskal-Wallis test among statistical groups as 0.05. For predicting the microbe function, the Greengenes database (version 13_8, 99 % OTU) and VSEARCH were applied for close-reference clustering, followed by PICRUSt to identify the corresponding function in the KEGG database.

### Quantification of ARGs

6 genes were quantified, including the 16S rRNA encoding gene and ARGs - *bla*_*TEM*_, *bla*_*CTX-M-32*_, *sul1, mcr-1*, and the *class 1 integron integrase intI1*. All qPCRs were performed in 20μl reaction volumes on a CFX96 Touch real-time PCR detection system (Bio-Rad, USA). First, a standard curve was determined using 5 serial dilutions with a known amount of each gene as a template. Standards in duplicates and samples in triplicates were analysed. Ct values were obtained and copy number calculations were done in Microsoft Excel. Graphs were plotted using GraphPad Prism. For normalisation and plotting, the copy number of each ARG was divided by the copy number of 16s rRNA genes.

## Results and Discussion

The twenty most prevalent genera are represented in Fig 1a. Community profile in terms of richness and evenness remained mostly consistent across all the samples and can be verified through various alpha diversity indices represented in Fig 1b. Interestingly, a distinct clustering observed in the beta-diversity index showed dissimilarity across months (PERMANOVA, R2 value=0.3965, p-value=0.001). This suggested microbial profiles fluctuated over the months regarding genera or their abundance. One biologically significant change between sampling timepoints that could have affected the profiles was the increase in COVID-19 cases.

**Fig 1.**
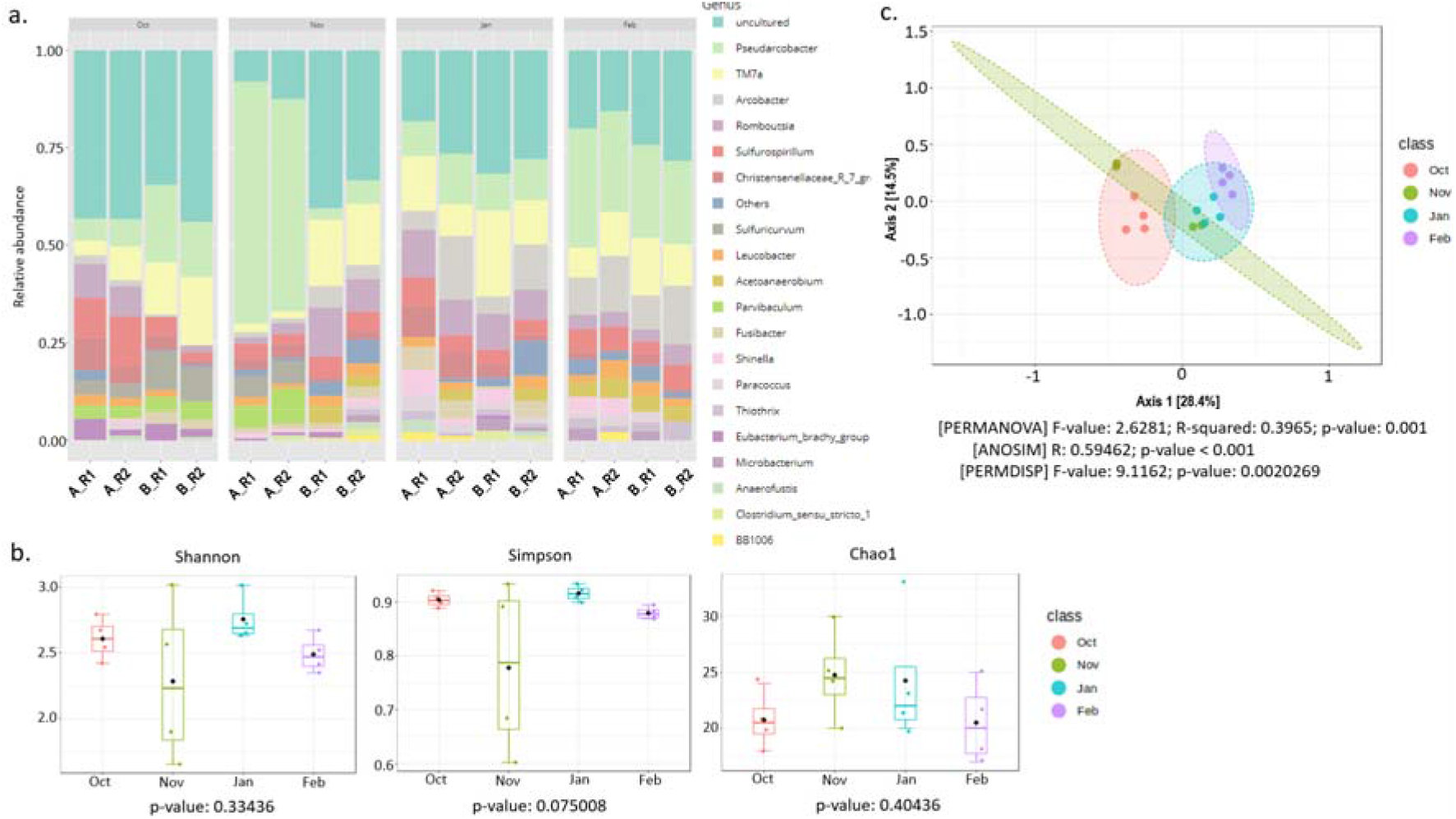
**a. Relative abundance of known bacterial genera:** Bar plot representing the prevalent genera across water samples. The colour scheme represents different taxa, as indicated by the legend on the right. **b. Richness and evenness within the samples:** The boxplots show alpha diversity calculated using the Shannon Index, Simpson and Chao1. The p-values >0.05 indicate nearly even and consistent diversity across the surveyed months. **c. Feature dissimilarity across samples:** The PcoA analysis using the Bray-Curtis dissimilarity index shows about 42.9% variation (a distinct clustering) of sample features across months (Permanova, p=0.001). Statistics from other beta diversity tests are also listed.

We examined the core and other prevalent members to find any possible correlation (Fig 2a and Fig 2b). Since October and November 2020 saw relatively lower cases (∼ 167 cases), they were classified as “low case incidence” months (less than 200 cases per month) and January and February 2021 were classified as “high case incidence” months (more than 1000 cases per month) as by then, the disease incidence had increased significantly (∼1780).

**Fig 2.**
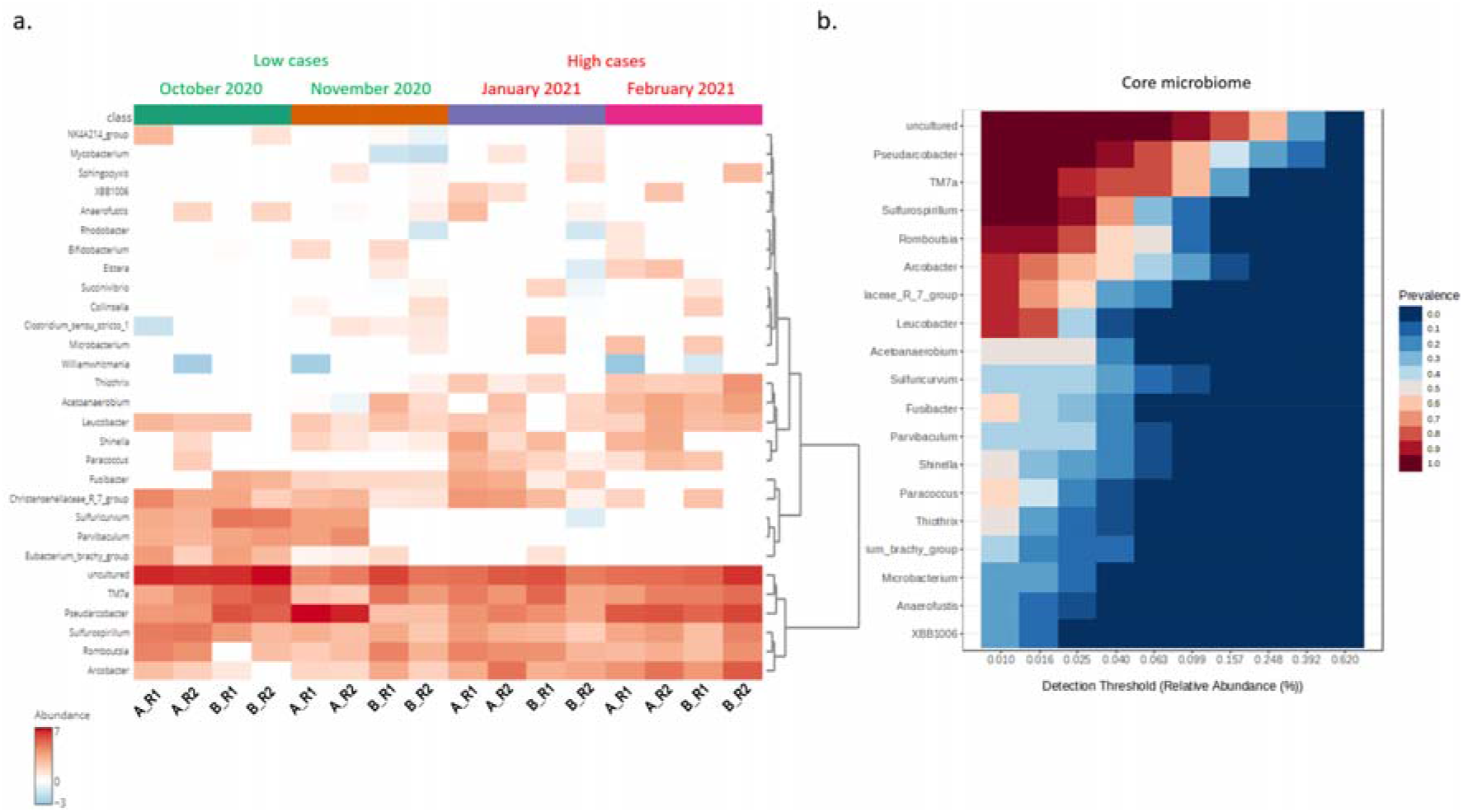
**a. Clustering analysis of the wastewater samples**. Heatmap and hierarchical clustering of the main genera associated with samples across all months. The heatmap shows the relative abundance of the top 20 bacterial genera (rows) in each sample (columns). The colour gradient indicates the range of the relative abundance. **b. Heatmap of the core microbiome analysis to identify core taxa at the genus level**. A total of (number) samples are included in the visual representation. The y-axis represents the prevalence level of core features across the detection threshold (relative abundance) range on the x-axis. Each genus’s prevalence variation is indicated by a colour gradient from blue (lowest) to red (highest).

We observed 7 core members, with the highest abundance attributed to uncultured microbes. The second most abundant genera were Pseudoarcobacter from the Arcobacteraceae family. They are found in raw sewage water and are associated with intestinal and extraintestinal diseases, abdominal pain, nausea, and fever (Kietsiri et al. 2021). Up next were TM7 from the Saccharimonadaceae family. They are also known to prevail in wastewater and are linked to gut and oral inflammation (Bor et al. 2019; Hugenholtz et al. 2001). The third highest abundance was Sulfurospirillum. Although not known as pathogenic, they thrive in polluted habitats as their substrate range largely involves toxic compounds such as arsenate (van den Berg et al. 2016). Next is Rombutsia, a newly discovered Gram-positive member of Peptostreptococcaceae. This family has established associations with colorectal cancer and several bowel diseases (Chang et al. 2023). Next was Arcobacter, which had similar implications to Pseudoarcobacters. Apart from the core members, other genera were enriched distinctively during either the high or low disease incidence months.

We performed LEfSe analysis to compare this differential abundance across months, categorised by low or high COVID-19 incidence. A histogram of the LDA scores (Fig 3a) indicates that the low case period was characterised by the prevalence of mostly uncultured genera along with common gut microbiota members like Erysipelotrichaceae (Shanks et al. 2013), *Sedinibacterium* which is commonly found in sewage sediments (Song et al. 2017) and Methanosarcinales found in landfill heaps and even animal gut (Volmer et al. 2023). Interestingly, various pathogenic genera were enriched during the high phase of COVID-19 infections. The most differentially abundant bacterial orders were Theotricales, Flavobacteriales, Rhodobacterales, Neisseriales, and Campylobacterales, amongst others (LDA score[log10]>4).

**Fig 3.**
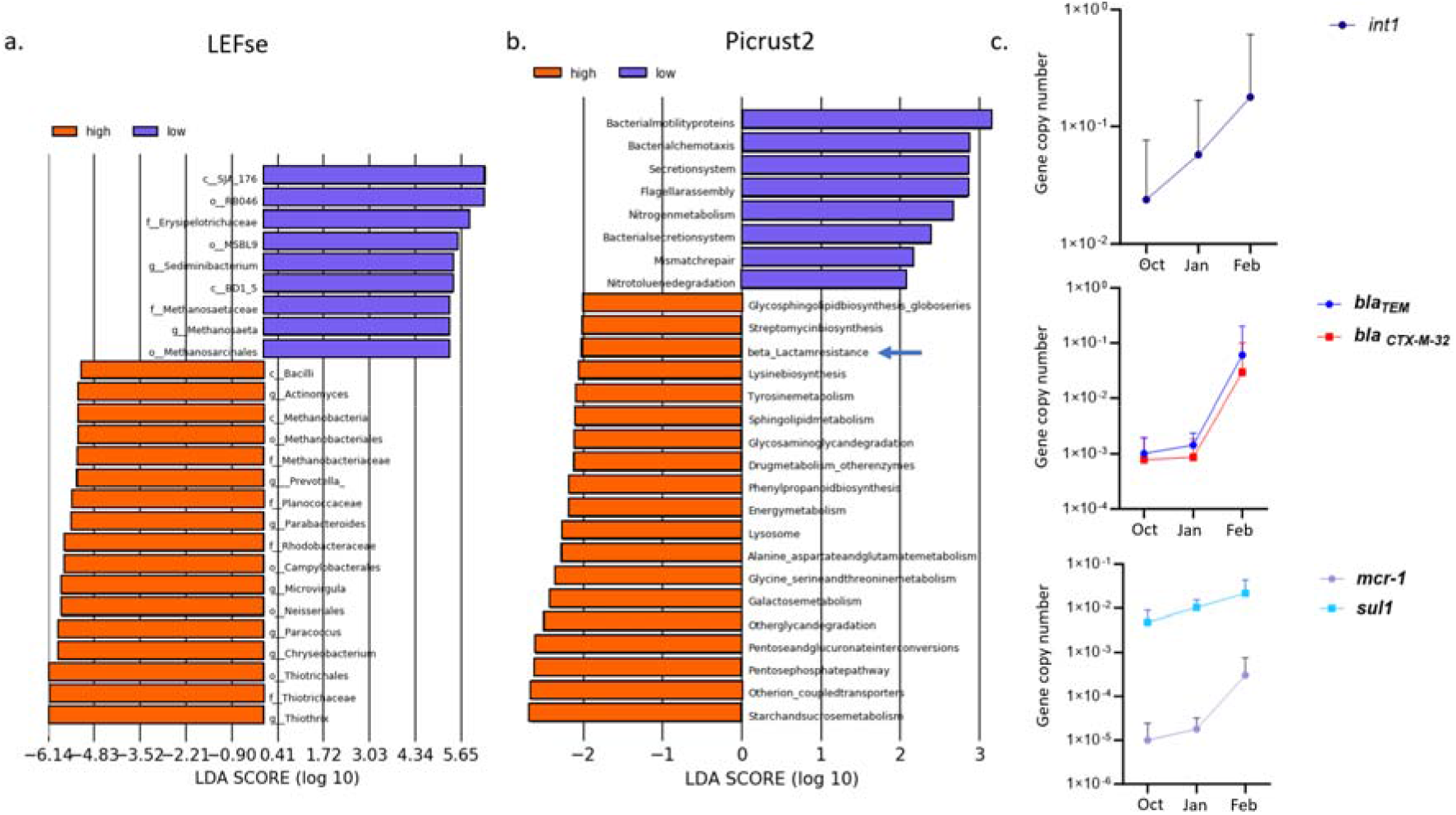
**a. LDA plot** revealing differentially abundant taxa with LDA score (log10) > 2 and P < 0.05 between high (red) and low case (green) samples. **b. predictive functional enrichment** due to various taxonomic groups (LDA score >2, P<0.05) in high (orange) and low (purple) case samples determined using PICRUST2. **c. The gene copy number** of various ARGs was normalised against the 16S rRNA gene copy number on a log-transformed scale.

We hypothesise that such differential abundance of taxa must have also led to a functional enrichment. The LDA score histogram in Fig 3b represents various functions that were distinctly enhanced. While both phases showed enrichment in classic metabolic and cellular pathways like chemotaxis, substrate synthesis & degradation, the high case period showed an intriguing enhancement of drug metabolism, specifically beta-lactams and aminoglycosides (LDA score (log10) > 2).

We verified this enrichment by determining the gene copy numbers of ARGs across samples of three different months. October 2020 represented the low case period, and the high case period represented January & February 2021. 6 genes including the 16S rRNA gene and ARGs - *blaTEM, blaCTX-M-32, sul1, mcr-1*, and the class 1 integron integrase *intI1* were quantified (Fig 3c). We found that copy numbers of beta-lactams (*blaTEM* and *blaCTX-M-32*) dramatically increased in February 2021, indicating widespread ESBL resistance amongst the bacterial population during the high case period. Other ARGs, such as *sul1* and *mcr-1*, also increased during this period. Mobile genetic element - *int1* was present and increased during the high case period. This could mean an extensive increase in the spread of ARGs during the outbreak (Johansson et al. 2023).

## Conclusion

This study utilised 16s rRNA metagenomic profiling of untreated wastewater to determine the microbial diversity around Klang Valley, Malaysia. While we acknowledge that species-level identification provides higher resolution data, genus-level 16S rRNA analysis remains a widely accepted and valuable approach in microbial community studies. The focus on genus-level identification in our study allows for the detection of significant patterns and trends within the microbial community, which are essential for understanding the overall ecosystem dynamics.

This study found that although the core member profile of the wastewater remained highly consistent during both phases of COVID-19, certain taxa were differentially abundant during the high-case periods. Alarmingly, the predominant core members (irrespective of the outbreak phase) were mostly pathogens like Arcobacters and Pseudoarcobacters -families whose members are known to cause bacteraemia and peritonitis, with some members classified as “serious hazard to humans” by International Commission on Microbiological Specification for Food (Kietsiri et al. 2021); TM7 known to cause oral inflammation (Hugenholtz et al. 2001); Peptostreptococcaceae member Rombutsia-a family associated with bowel diseases and colorectal cancer (Chang et al. 2023). Exposure to these pathogens through treated/untreated water could harm human and animal health. Interestingly, Thiotricales and Flavobacteriales were also enriched in the high case periods, possibly due to the surge in sulphur-based soaps and detergents from the standard protective measurements implemented during the COVID-19 pandemic (Cheng et al. 2024). In addition, Neisseriales and Campylobacterales – contaminants of human faecal matter-could have been enriched by static population density around the region due to the travel restrictions under the Movement Control Order (MCO) that was in effect from March 2020 to November 2021 in Kuala Lumpur. Our functional enrichment analysis showed an increased spectinomycin synthesis pathway, indicating the prevalence of spectinomycin-producing bacteria and genes. A spike in their numbers could be due to increased antibiotic precursors in waste discharged from hospitals and residential areas during high case periods, which were easily available for the bacteria (Davies and Davies 2010) This may also have contributed to the dissemination of ARGs through horizontal gene transfer events and is supported by the increased *int1* copy number (Fig 3c). ARG profiling of these samples showed the prevalence of *sul1* and *mcr-1*, which could mean a surge in sulphonamide and colistin drug resistance, apart from the more widespread Extended-spectrum beta-lactamases (ESBL) resistance. The prevalence of mobile elements is particularly concerning since they could mean an ever-increasing transfer of ARGs across species and hence may lead to a surge of multi-drug resistance in previously susceptible strains, especially in a post-pandemic era when sub-lethal concentrations of antibiotics are quite common in wastewater.

This study is intended as a preliminary investigation to explore the potential impact of COVID-19 on wastewater microbial communities. The constraints of the pandemic, including logistical challenges and limited access to resources, necessitated a smaller sample size for this initial study. Despite these limitations, we believe that our findings provide valuable preliminary insights and highlight the need for more extensive future research. Prospective studies should aim to include a greater number of samples and account for a broader range of environmental and operational parameters to validate and build upon our initial observations.

## Acknowledgements

We are thankful to Indah Water Konsortium Sdn. Bhd for allowing and assisting in sample collection. We acknowledge Monash University Malaysia Genomics Platform (MUMGP), the School of Science, Monash University, for providing the required infrastructure for this research activity. We extend our gratitude to Muhammad Zarul Hanifah Md Zoqratt, supported by the Monash Malaysia R&D Sdn. BhD. (MMR&D) and others who directly or indirectly assisted in this study.

## Funding

This research received no specific grant from funding agencies in the public, commercial, or not-for-profit sectors. MUMGP was supported by a core grant from the School of Science.

## Conflict of Interests

The authors report there are no competing interests to declare.

## Author Contributions

US wrote the manuscript and performed the experiments; HSL and LSY performed the experiments and data analysis; AMA and KBC provided access to the facility and collected samples; QA provided access to the sequencing facility and reviewed the manuscript; THS conceptualise the study, provide supervision, funding, data analysis and review the manuscript.

## Data Availability

The 16S rRNA sequence data that support the findings of this study have been deposited in the Sequence Read Archive (SRA) with the primary accession code <u>PRJNA1095152</u>

## Ethics Approval

There were no ethical approvals required for this study.

## Notes

### Competing Interest Statement

The authors have declared no competing interest.

